# Meta-analysis on safety of standard vs prolonged infusion of beta-lactams

**DOI:** 10.1101/2024.04.08.24305493

**Authors:** Hunter Rolain, Zachary Schwartz, Raymond Jubriail, Kevin Downes, Lisa Hong, Alireza Fakhri Ravari, Nathaniel J. Rhodes, Marc H. Scheetz

**Affiliations:** Midwestern University-Downers Grove Campus, Chicago College of Osteopathic Medicine, Downers Grove, IL, USA; Midwestern University-Downers Grove Campus, Department of Pharmacy Practice, Downers Grove, IL, USA; Midwestern University-Downers Grove Campus, Pharmacometrics Center of Excellence, Downers Grove, IL, USA; Northwestern Memorial Hospital, Department of Pharmacy, Chicago, IL, USA; Division of Infectious Diseases, Children’s Hospital of Philadelphia, Philadelphia, PA, USA; Department of Pediatrics, Perelman School of Medicine of the University of Pennsylvania, Philadelphia, PA, USA; Loma Linda University School of Pharmacy, Department of Pharmacy Practice, Loma Linda, CA, USA

**Keywords:** standard infusion, prolonged infusion, renal failure, beta-lactams, kidney damage, neurotoxicity, kidney injury, cefepime, piperacillin, ceftriaxone

## Abstract

**Background:** Efficacy for prolonged infusion beta-lactam dosing schemes has been previously described, but there has been less focus on the safety of standard vs prolonged infusion protocols of beta-lactams. This study explored differences in adverse drug reactions (ADRs) reported for beta-lactams between each of these infusion protocols.

**Methods:** A systematic review of MEDLINE literature databases via PubMed was conducted and references were compiled. Articles were compiled and assessed with specific inclusion/exclusion criteria. We included randomized and nonrandomized, prospective, and retrospective cohort studies that reported adverse effects due to either standard (30-60 mins) or prolonged (≥3 hours) infusions of beta-lactam infusions. Total ADRs between strategies were analyzed by infusion methodology. The most consistently reported ADRs were subject to meta-analysis across studies.

**Results:** 13 studies met inclusion/exclusion criteria with data for 4184 patients. There was insufficient data to systematically analyze neurotoxicity or cytopenias. Eight studies reported on nephrotoxicity outcomes with no significant difference in event rates between standard (n=440/2117, 20.8%) vs prolonged infusion (n=264/1284, 20.6%) of beta-lactams (OR=1.09, 95% CI [0.91, 1.30]). Six studies reported on rates of diarrhea with no significant difference in event rates between standard (n=21/359, 5.8%) vs prolonged infusion (n=25/330, 7.6%) of beta-lactams (OR=1.33, 95% CI [0.71,2.47).

**Conclusion:** Prolonged and standard infusion schemes for beta-lactams demonstrated adverse effects at similar rates for both infusion schemes. Future research should focus on improved standardization of adverse effect definitions and a priori aim to study neurotoxicity and cytopenias. Consistent recording of ADRs and standardized definitions of these reactions will be paramount to further study of this subject.

## Background

Beta-lactams are among the most frequently utilized antibiotics worldwide [1,2] as they are first line options for multiple infectious syndromes [1-4]. Among the high use beta-lactam antibiotics are broad-spectrum cephalosporins such as cefepime, penicillin-beta-lactamase inhibitor combinations such as piperacillin-tazobactam, and carbapenems such as meropenem. Most often, beta-lactams are delivered as intermittent infusions (IIs) over a period of time ranging from 30 to 60 minutes (i.e., standard infusion (SI)), typically as a 30-minute infusion [5]. Because bacterial killing is improved when time over bacterial minimum inhibitory concentrations (MICs) is optimized, beta-lactams are increasingly being utilized in prolonged infusion platforms to improve efficacy [1-4, 6-17]. Prolonged infusions (PI) consist of extended infusions (EIs) that are longer than standard infusions (e.g., 3-4 hours) and continuous infusions (CI) that deliver constant concentrations of antibiotics without plans to stop. In this study, we classified both EI and CI as PI.

The efficacy of prolonged infusions has been shown to be either improved or equal to standard infusions [5,18-20,21]. Safety, however, has been less well explored. As with many drugs, ADRs for beta-lactams do occur, albeit infrequently, and are usually mild to moderate in severity. ADRs driven by dose and exposure are of particular interest to clinicians when a biologic relationship is identified, as they can be predicted and avoided. One such example of dose-dependent toxicity in beta-lactams is neurotoxicity [22,23]. Nephrotoxicity, on the other hand, is generally thought to be dose-independent and related to allergic-mediated mechanisms and type-II hypersensitivity reactions [22]. Although dose-toxicity relationships are reasonably established, the time course in which the dose is delivered has been less studied. When beta-lactams are given as standard infusions, higher serum and tissue peak concentrations are obtained while troughs are lower. In contrast, when prolonged infusions are utilized, peaks are lower while trough levels are higher. In both of these infusion types, for a fixed dose, the area under the curve (AUC, or total exposure) is the same. It is unknown if differences in pharmacokinetic parameters (e.g., different Cmax yet similar AUC) lead to differences in ADRs. Because there are few large and purpose-defined trials comparing the safety of standard versus prolonged infusion methods for the assessment of safety, we performed a systematic review to identify studies that documented rates of ADRs in PI and SI groups. Meta-analyses were performed for the most consistently reported ADRs between the groups.

## Methods

### Objective

The primary objective for this meta-analysis was to determine the difference in incidence of ADRs in standard versus prolonged infusions of beta-lactams.

### Search Strategy and Data Collection

To begin, all literature that was previously identified by the international consensus recommendations [24] under PICO question 7, “Is use of a prolonged-infusion beta-lactam safer than standard infusion in children” was considered. Then an updated literature search was completed using PubMed and included the following key search terms: “standard infusion,” “extended infusion,” “renal failure,” “kidney damage,” “kidney injury,” “neurotoxicity,” “beta-lactam,” “cefepime,” “piperacillin,” “ceftriaxone.” All articles were compiled after review of an inclusion/exclusion criteria in order to determine eligibility.

### Inclusion/Exclusion Criteria

To be eligible, studies were required to meet the following criteria: 1) randomized trials or non-randomized, prospective, or retrospective cohort studies; 2) ADRs reported; 3) beta-lactam usage either alone or in combination with other antibiotics consistent across both treatment arms; and 4) only 2 treatment arms (i.e., standard infusion and prolonged infusion). Studies were excluded for concomitant drug use varying amongst treatment arms, mixed use of standard infusion and prolonged infusion at the individual patient level, or not meeting inclusion criteria.

### Data Extraction

Pertinent data was entered into a data extraction table (Table 1). The data listed was collected for each individual study: studies characteristics (authors, publication year), patient population (age, disease state), treatment regimens (specific beta-lactam, infusion type), and ADRs within each group.

**Table 1.** Data Extraction Table.

| Study | Year | Total Patient<br>s | # of<br>Subjects<br>of EI | # of<br>ADRs in<br>EI | % of<br>ADRs in<br>EI | # of<br>Subjects<br>in SI | # of<br>ADRs in<br>Standard<br>Infusion | % of<br>ADRs in<br>SI | Study Design |
| --- | --- | --- | --- | --- | --- | --- | --- | --- | --- |
| Bauer<sup>8</sup> | 2013 | 132 | 83 | 25 | 30.1% | 49 | 12 | 24.5% | Retrospective |
| Abdul-Aziz<sup>9</sup> | 2016 | 140 | 70 | 0 | 0% | 70 | 0 | 0% | RCT |
| Chytra<sup>10</sup> | 2012 | 240 | 120 | 10 | 8.3% | 120 | 12 | 10% | RCT |
| Grant<sup>41</sup> | 2002 | 98 | 47 | 0 | 0% | 51 | 0 | 0% | RCT |
| Bao<sup>33</sup> | 2017 | 50 | 25 | 11 | 44% | 25 | 16 | 64% | RCT |
| Fan<sup>42</sup> | 2017 | 367 | 182 | 0 | 0% | 185 | 0 | 0% | RCT |
| McCormick<sup>43</sup> | 2015 | 200 | 100 | 9 | 9% | 100 | 11 | 11% | Retrospective |
| Mousavi<sup>44</sup> | 2017 | 272 | 136 | 45 | 32.9% | 136 | 40 | 29.3% | Retrospective |
| Dulhunty<sup>45</sup> | 2013 | 60 | 30 | 0 | 0% | 30 | 0 | 0% | RCT |
| Nicolau<sup>46</sup> | 2001 | 35 | 17 | 11 | 64.7% | 10 | 9 | 70% | RCT |
| Contrina-Luque<sup>47</sup> | 2016 | 78 | 40 | 0 | 0% | 38 | 0 | 0% | RCT |
| Van Zanten<sup>13</sup> | 2006 | 93 | 47 | 0 | 0% | 46 | 0 | 0% | RCT |
| Shabaan<sup>14</sup> | 2017 | 102 | 51 | 12 | 23.5% | 51 | 29 | 56.9% | RCT |
| McNabb<sup>36</sup> | 2001 | 35 | 17 | 9 | 52.9% | 18 | 13 | 72.2% | RCT |
| Karino<sup>48</sup> | 2016 | 320 | 160 | 52 | 32.5% | 160 | 53 | 33.1% | Retrospective |
| Winstead<sup>49</sup> | 2016 | 181 | 86 | 0 | 0% | 95 | 0 | 0% | Retrospective |
| Knoderer<sup>15*</sup> | 2017 | 50 | 50 | 0 | 0% | 0 | 0 | 0% | Retrospective |
| Ram<sup>16</sup> | 2018 | 91 | 47 | 3 | 6.4% | 58 | 6 | 6.9% | RCT |
| Nichols<sup>50</sup> | 2015 | 150 | 143 | 0 | 0% | 7 | 0 | 0% | Retrospective |
| Cotner<sup>51</sup> | 2017 | 2390 | 690 | 149 | 21.6% | 1700 | 316 | 18.6% | Retrospective |
| Roberts<sup>20</sup> | 2006 | 57 | 28 | 0 | 0% | 29 | 0 | 0% | RCT |
| Schmees<sup>52</sup> | 2016 | 113 | 61 | 0 | 0% | 52 | 0 | 0% | Retrospective |
| Nichols<sup>31</sup> | 2019 | 67 | 21 | 2 | 14.3% | 46 | 3 | 6.5% | Retrospective |
| Lau<sup>30</sup> | 2006 | 262 | 130 | 14 | 10.8% | 132 | 16 | 12.1% | RCT |
| Padari<sup>17</sup> | 2012 | 19 | 10 | 0 | 0% | 9 | 0 | 0% | RCT |
| Monti<sup>21</sup> | 2023 | 607 | 303 | 0 | 0% | 304 | 0 | 0% | RCT |

\* Insufficient adverse events recording

### Data Analysis

Meta-analyses were performed for ADRs that occurred in 5 or more studies only. Statistics for the meta-analysis was performed using Review Manager V. 5.4 from Cochrane Library [25]. Fixed Odd-Ratios were calculated from the dichotomously reported ADR rates from each study with 95% Confidence Interval in RevMan [25]. Analysis and forest plot summary of the pooled adverse events were created in RevMan [25] (Figure 2 & 3). The quality of evidence and risk for bias was independently assessed by 3 study authors with majority rule for final classification. Analysis of study quality was performed via Newcastle-Ottawa Scale [26] (Table 3) and Cochrane Risk Bias [27] (Table 4) for retrospective and prospective studies, respectively. Publication bias was assessed visually using funnel plot inspection for nephrotoxicity (Figure 4) and diarrhea (Figure 5).

## Results

After initial screening, a total of 26 studies were assessed for eligibility for meta-analysis based on title and abstract (Figure 1). Nephrotoxicity and diarrhea were the most prevalent reported ADRs according to the inclusion criteria for the meta-analysis. Out of the 26 studies, 12 were excluded because no ADRs were reported (Figure 1). One study was excluded due to insufficient adverse events recording (Figure 1). After exclusion, a total of seven randomized control trials and six retrospective studies met inclusion criteria with data for 4196 patients. Eight studies reported nephrotoxicity from a total of 3564 patients with no significant difference in patients of standard (n=436/2276, 19.8%) vs prolonged infusion (n=267/1288, 20.6%) of beta-lactams (OR=1.08, 95% Confidence Interval [0.91, 1.29]) (Figure 2). Among the eight studies, five of them reported concomitant administration of vancomycin, 4 reported the exact number of patients receiving it. The total number of patients receiving concomitant vancomycin in these studies was 80.3%. Seven studies observed diarrhea in a total of 794 patients with no significant difference in patients of standard (n=21/417, 5.0%) vs prolonged (n=25/377, 6.6%) infusion of beta-lactams (OR=1.30, 95% Confidence Interval [0.71,2.37]) (Figure 3). No publication bias was detected for either nephrotoxicity (Figure 4) or diarrhea (Figure 5).

**Figure 1:**
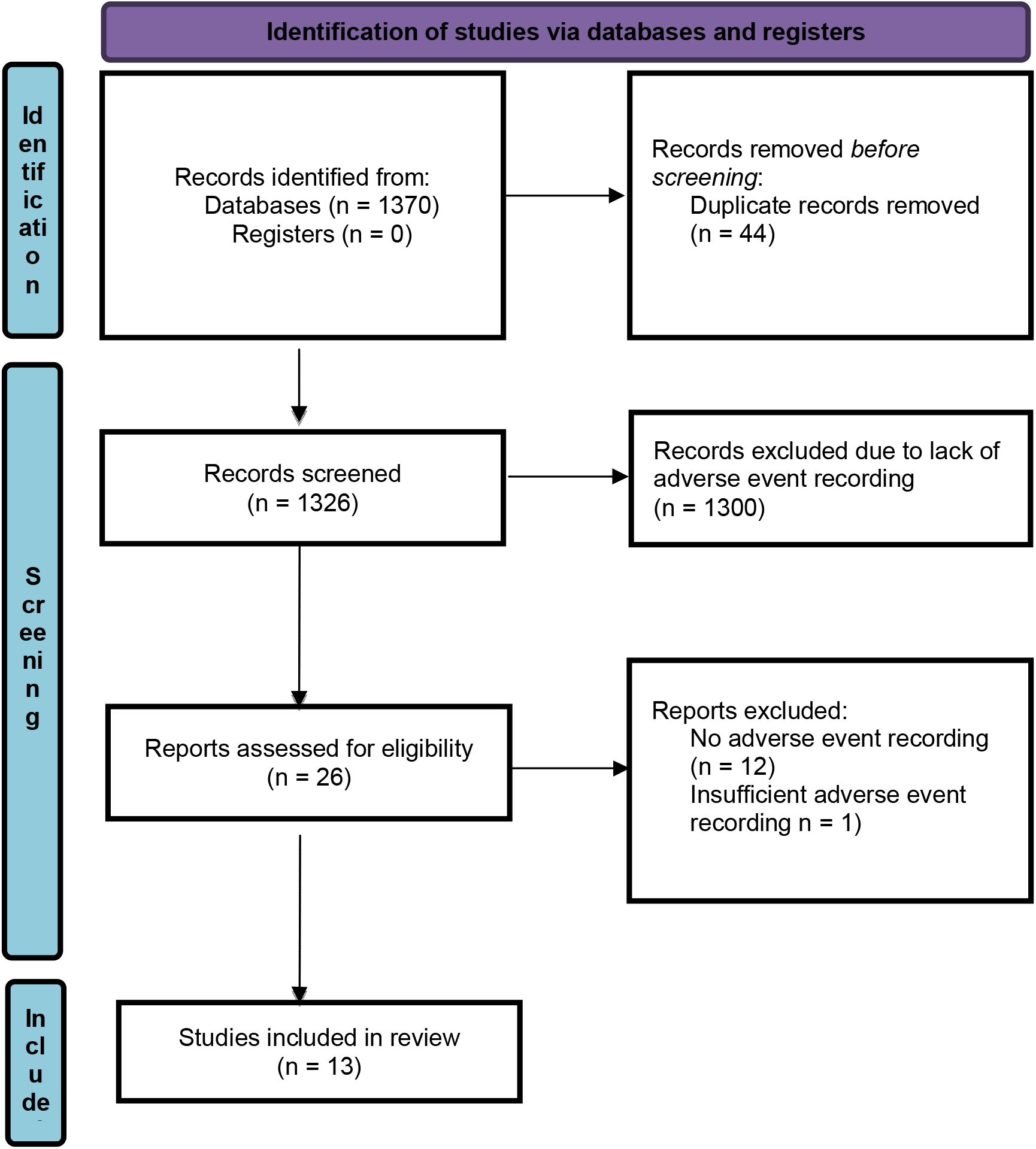
PRSIMA 2020 Flow Diagram for research identification

**Figure 2:**
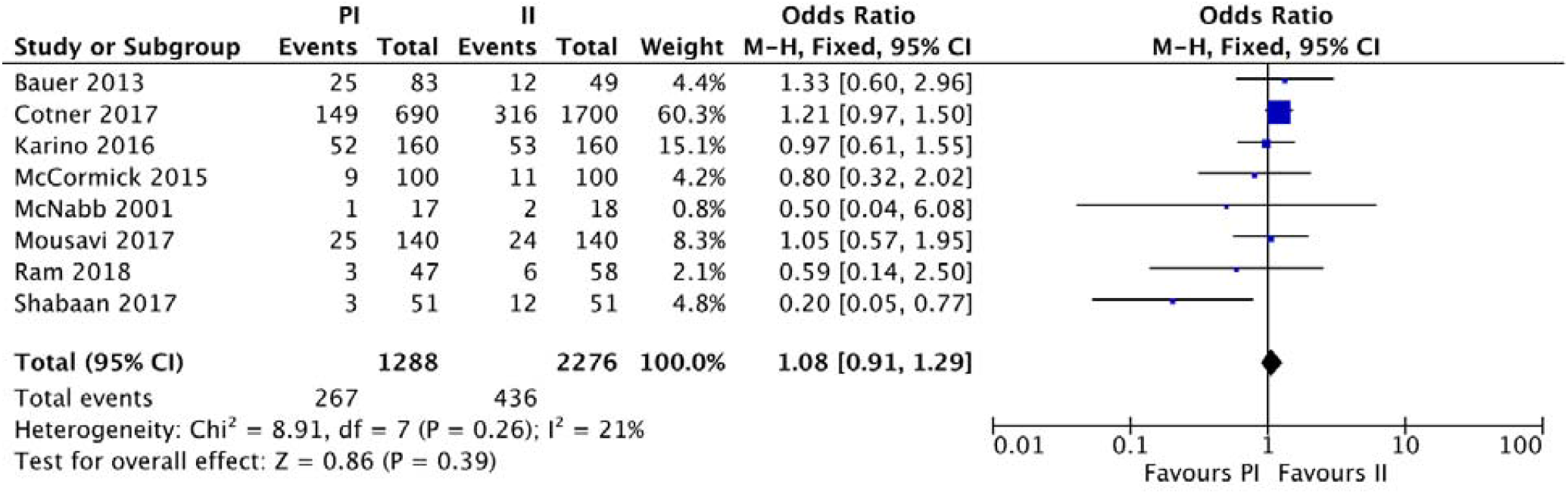
Forest plot of the adverse side reaction of nephrotoxicity from prolonged (PI) vs standard infusion (II) of beta-lactams

**Figure 3:**
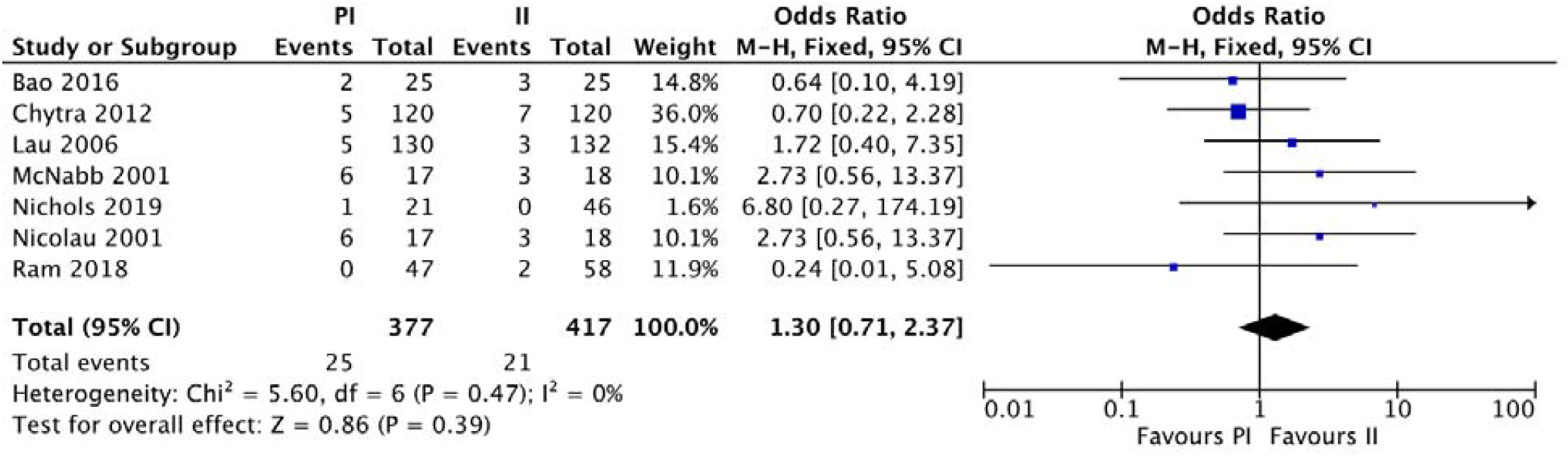
Forest plot of the adverse side reaction of diarrhea from prolonged (PI) vs standard infusion (II) of beta-lactams

**Figure 4.**
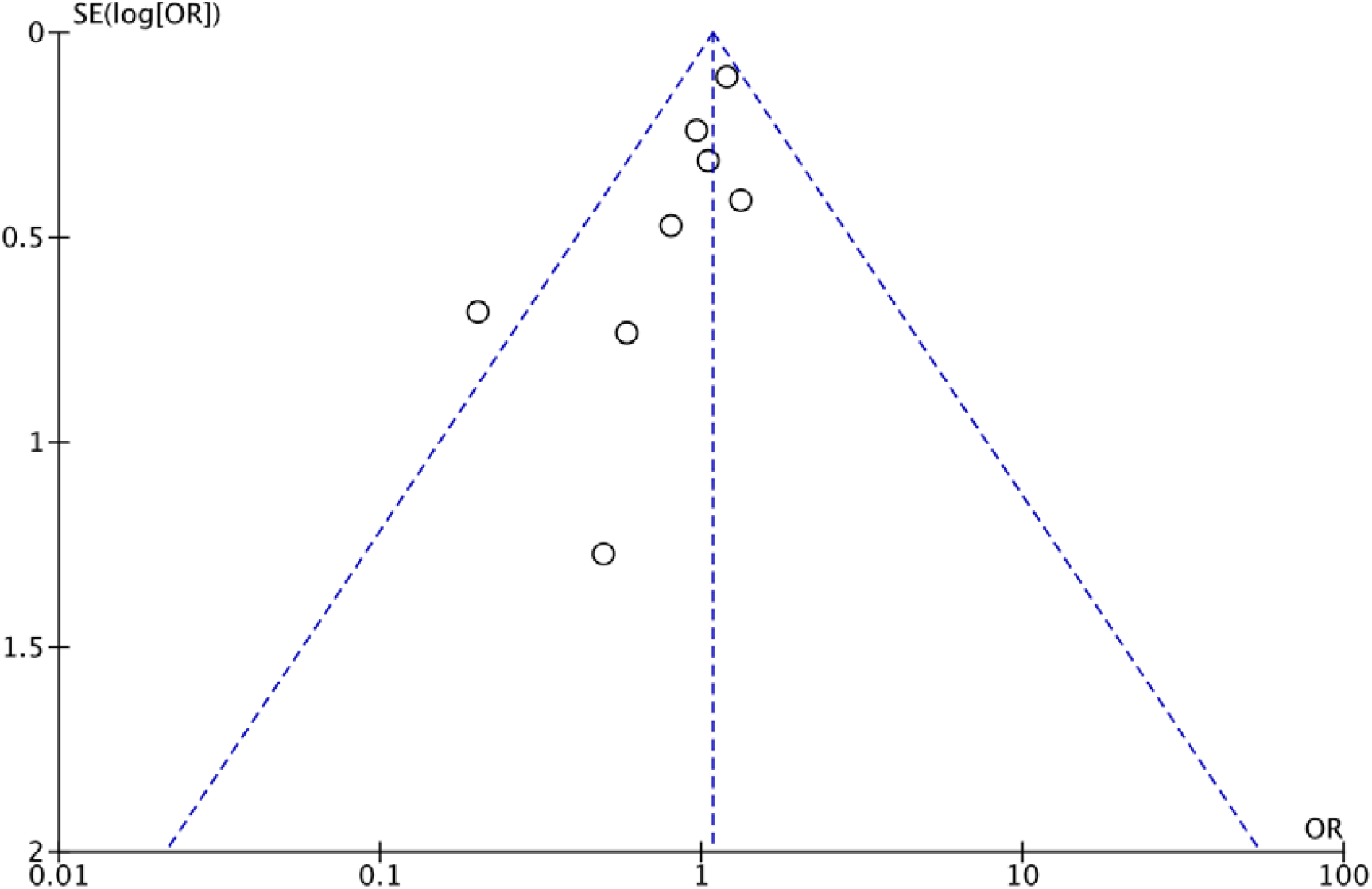
Funnel plot of the adverse side reaction of nephrotoxicity from prolonged (PI) vs standard infusion (II) of beta-lactams

**Figure 5.**
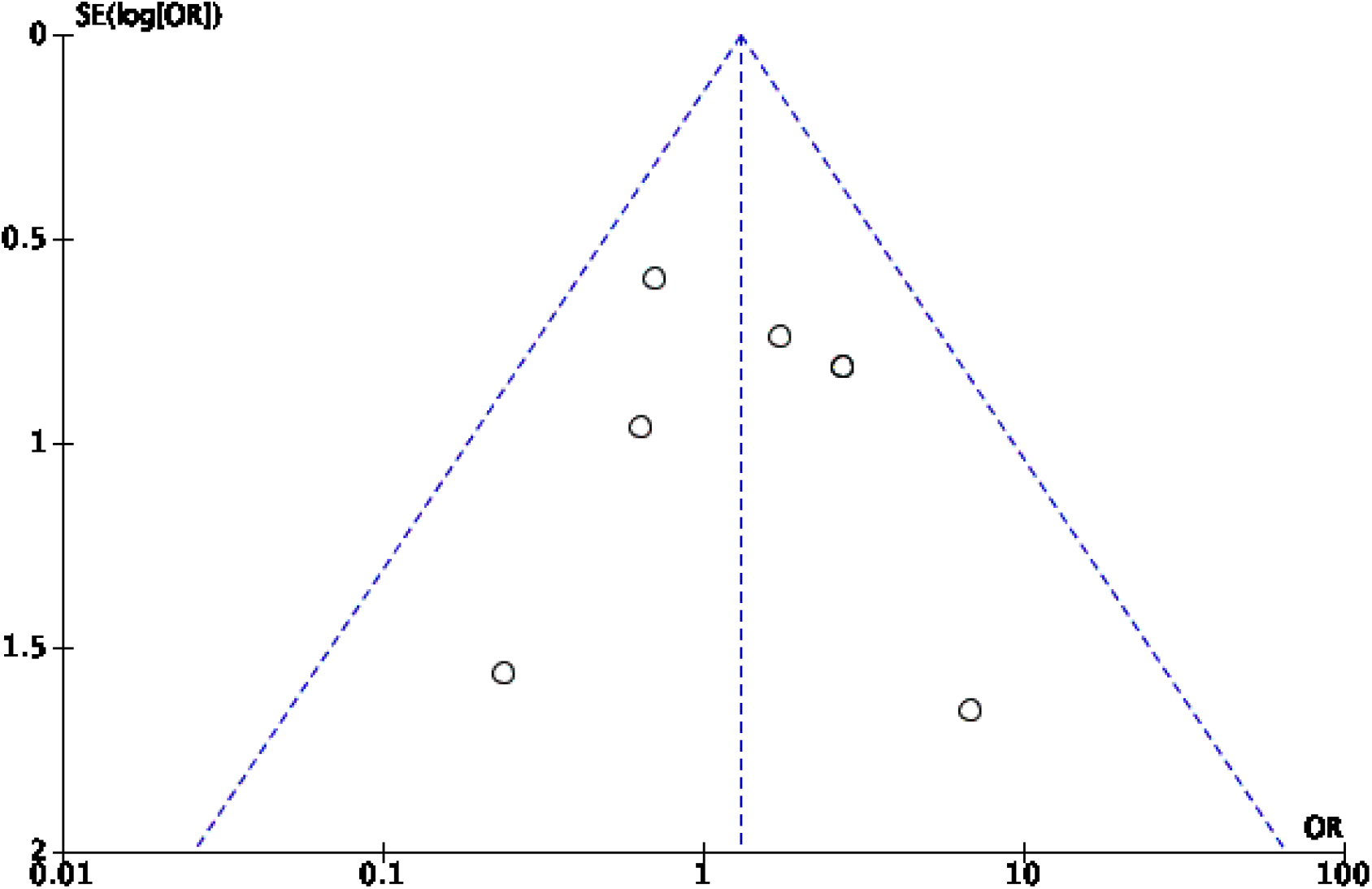
Funnel plot of the adverse side reaction of diarrhea from prolonged (PI) vs standard infusion (II) of beta-lactams

## Discussion

Our review and meta-analysis did not identify a signal for differences in ADRs between prolonged and standard infusion schemes. Lack of a safety signal is an important finding, given that prolonged infusions are being increasingly used to improve efficacy. Thus, if future studies are to reach the same conclusion, the most efficacious infusion protocols can be used without additional safety concerns.

Beta-lactams are generally regarded as safe agents, associated with minimal adverse effects. Nephrotoxicity and diarrhea were the ADRs most commonly recorded by the studies in question and were thus studied via meta-analysis. Other ADRs including cytopenias, neurotoxicity, electrolyte imbalance, elevated liver function tests, and rash were reported inconsistently, and it was thus not possible to quantitatively analyze these ADRs. Nephrotoxicity was classified broadly and as reported by the individual study (Table 2), but no difference was found between infusion strategies when the studies were pooled categorically. In future work, standardizing classification schema to acceptable standards (e.g., KDIGO) will result in more meaningful comparison of ADRs across studies. A single standard would help increase specificity. In the present analysis, while many of the studies reported nephrotoxicity, some used the Acute Kidney Injury Network (AKIN) criteria [28], others used the Risk, Injury, Failure, Loss of kidney function, and End-stage kidney disease (RIFLE) criteria [28], and one relied on independent nephrologist assessment. It is also important to note that kidney injury rates here of ∼20% are very unlikely due to the beta-lactam and are probably more representative of the severity of illness of patients for which beta-lactams are required (i.e. infectious syndromes). Patients often had concomitant medications that are known nephrotoxins (e.g. vancomycin, aminoglycosides). We did not quantify these concomitant medications since they were not consistently reported in a manner that facilitated quantification.

**Table 2.**
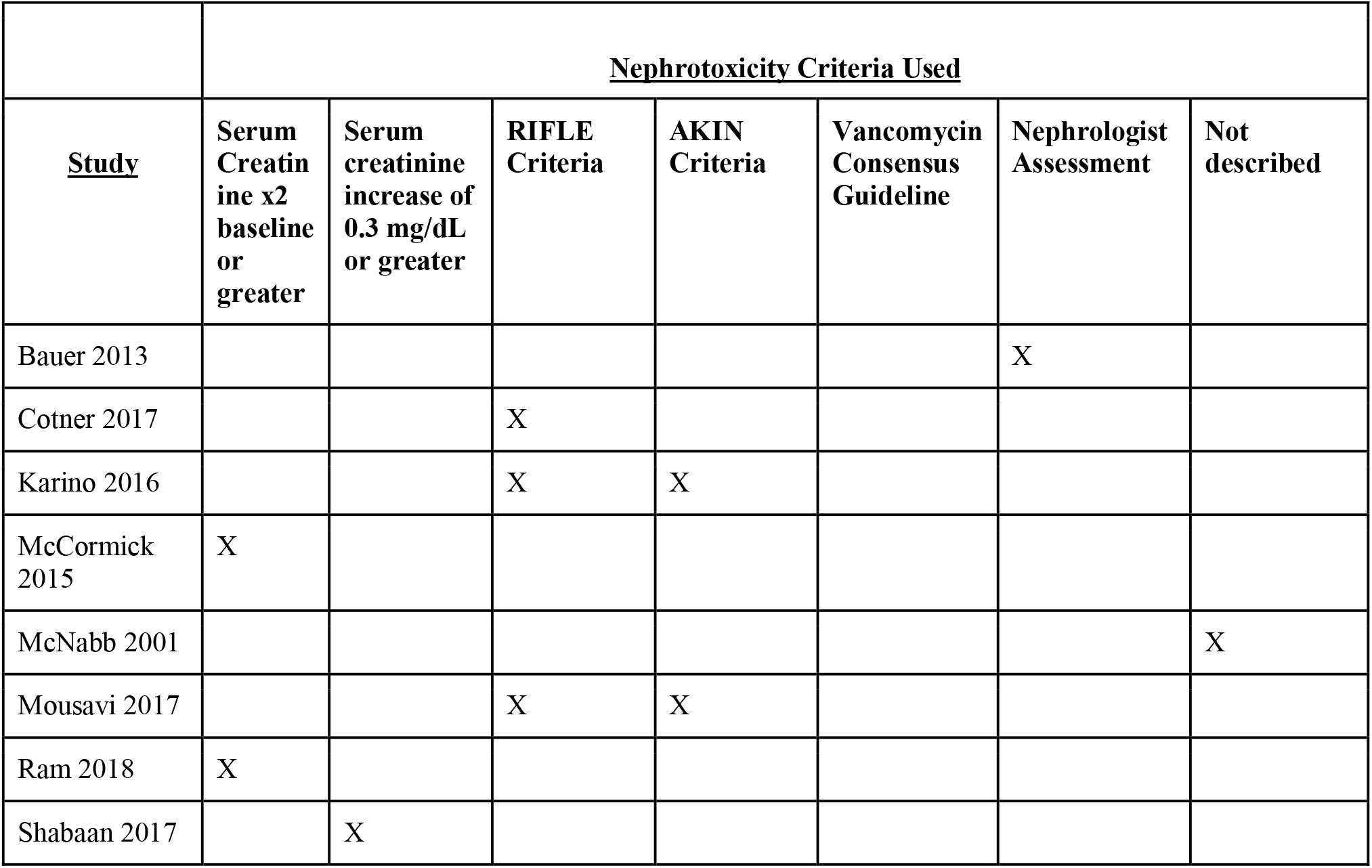
Data Extraction Table.

**Table 3.** Cochranes Risk of bias summary, review of author’s judgements about each risk of bias item for each included study.

| Study | Sequence Generation | Allocation concealment | Blinding | Incomplete outcome | Selective Reporting | Other bias | Intention to treat | Sample calculation |
| --- | --- | --- | --- | --- | --- | --- | --- | --- |
| McNabb 2001 | + | + | - | - | - | - | + | - |
| Ram 2018 | + | + | - | - | - | - | + | + |
| Shabbaan 2017 | + | + | + | - | - | - | + | + |
| Chytra 2012 | + | + | - | - | - | - | + | + |
| Bao 2017 | + | + | - | - | - | - | + | - |
| Nicolau 2001 | + | + | - | - | - | - | + | - |
| Lau 2006 | + | + | - | - | - | - | + | + |

+ Low Risk of Bias
- High Risk of Bias

**Table 4.**
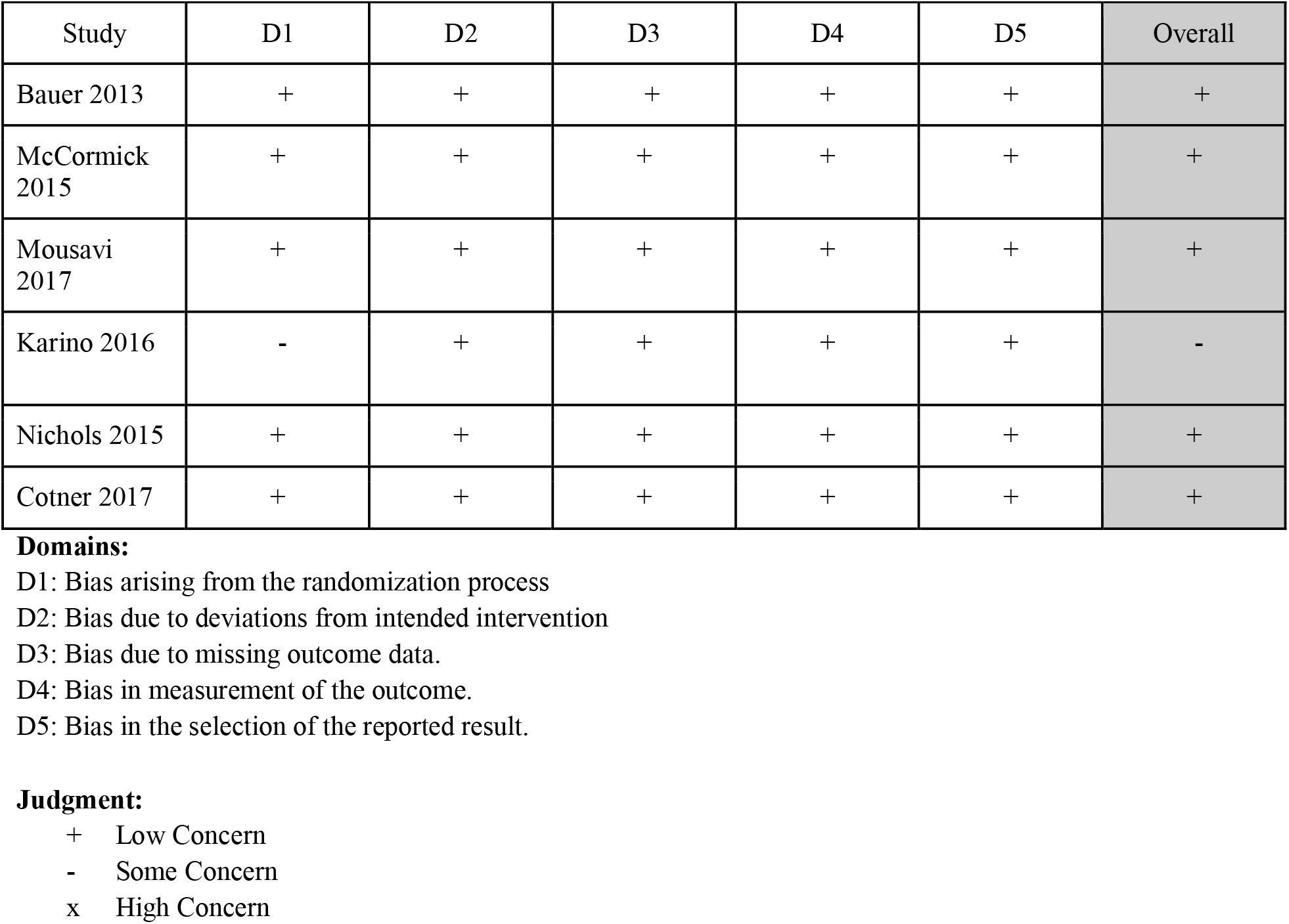
Risk of bias assessment using Newcastle-Ottawa Score.

Beta-lactam induced kidney injury can be separated into acute interstitial nephritis and direct cellular toxicity (e.g. acute tubular necrosis) with the former more common via an immune-mediated, dose-independent response. While the exact mechanism of drug induced acute interstitial nephritis has not entirely been elucidated, it is thought that the beta-lactam triggers an immune response by acting as a hapten, causing the drug to be immunogenic and subsequently upregulate immunoglobulins. These upregulated immunoglobulins result in an immune response mediated by CD8+ cytotoxic T-cells [29,30]. The destruction induced by these T-cells leads to tubular injury/acute kidney injury and ultimately renal failure in some cases [30]. Such a mechanism is thought to be dose and exposure independent. For certain beta-lactams such as imipenem, dose/exposure mediated events can occur and lead to cellular accumulation and mitochondrial stress; [31] however, in the case of imipenem, the addition of cilastatin is specifically used to prevent accumulation.

Diarrhea associated with beta-lactam usage is thought to be linked to drug concentration intensity and total time of therapy [10,12,32-36]. Diarrhea from antibiotics such as beta-lactams is multifactorial and can be caused by specific superinfection or more general gut microbiota dysbiosis. *Clostridioides difficile* infection is an example of superinfection, although the more common ‘antibiotic diarrhea’ is likely due to dysbiosis. We saw no difference between beta-lactam infusion strategies for any diarrhea reported.

Beta-lactam-induced neurotoxicity is proposed to be dose and exposure mediated [22,23,37,38].While it is less clear if maximal concentrations or total exposure drives the toxicity, administration of prolonged infusion beta-lactams which utilize lower or equal doses to standard intermittent infusions is hypothesized to result in less neurotoxicity [39,40]. Given that none of the studies comparing PI to SI antibiotics focused on sensitive methods that could monitor neurological findings (e.g. prespecified EEG testing), and that neurotoxicity is not a commonly reported ADR in general, it was not surprising that neurotoxicity was infrequently documented. Other adverse effects attributed to beta-lactams include rash, hepatic injury, and electrolyte imbalance. However, none of these ADRs occurred with considerable frequency in either group or were not measured in some of the studies, thus there was not enough data to quantitatively analyze them.

Limitations to this work include the frequent underestimation of ADRs in retrospective studies. Additionally, studies with subjects receiving both standard and prolonged infusions have not been specifically designed to detect differences in safety. As such, future studies should specifically place a focus on analyzing toxicity outcomes in a standardized manner for each of these infusion methods. It is important to also note that our meta-analysis utilized a definition of study-reported nephrotoxicity rather than a common unified definition. It was not possible to reclassify patients because the data were not available to do so. The definitions of nephrotoxicity, as presented in the studies, had considerable variation, with some studies lacking a clear and consistent characterization. Additionally, some of the studies included patients receiving concomitant nephrotoxic medications that may present a confounding variable. Cotner et. al, a study from which 2390 patients were included in this analysis, was a large retrospective study of hospitalized patients [51]. Many patients analyzed in the study required additional treatment with various nephrotoxic non-beta-lactam medications such as vancomycin (frequently administered with piperacillin/tazobactam), aminoglycosides, and loop diuretics that contributed to higher nephrotoxicity rates [51]. Finally, the characterization of ADRs in the majority of these studies were done in a clinical manner and was often left to physician characterization. Such reporting is subjective in nature. For future comparative studies between prolonged and standard infusion beta-lactams, it would be beneficial to utilize a standardized criteria for the classification of ADRs. An example includes the Common Terminology Criteria for Adverse Events (CTCAE) which is employed by organizations such as the National Cancer Institute and the FDA for this purpose. By utilizing an accepted standard set of criteria, the collection and utilization of safety data can be significantly improved. This will also result in enhanced external use of safety data for future research endeavors. In the context of our study, the incorporation of standardized criteria in future studies will yield the higher quality data that is required for a more comprehensive evaluation of the safety profile of these infusion protocols.

## Conclusion

Based on the dosing scheme of the analyzed studies, prolonged and standard beta-lactam infusion schemes displayed ADRs at similar rates. The most prevalent ADRs included nephrotoxicity and diarrhea; however, no difference was seen between PI and SI infusion strategies. Other ADRs were reported, however, frequency was low and reporting inconsistent. Further studies should be specifically designed to analyze toxicity outcomes from each of the infusion methods.

## Data Availability

Data that support the findings of this study are available from the corresponding author upon reasonable request. Data sharing will be subject to standard Data Use Agreements from Midwestern University.

